# Universal coordinate for cross-subject comparison of cardiovascular waveform dynamic on phase manifold

**DOI:** 10.1101/2024.09.09.24313272

**Authors:** Yu-Ting Lin, Ruey-Hsing Chou, Shen-Chih Wang, Cheng-Hsi Chang, Hau-Tieng Wu

**Affiliations:** Department of Anesthesiology, Taipei Veterans General Hospital, Taipei, Taiwan; School of Medicine, National Yang Ming Chiao Tung University, Taipei, Taiwan; Department of Critical Care Medicine, Taipei Veterans General Hospital, Taipei, Taiwan; Department of Anesthesiology, Shin Kong Wu Ho-Su Memorial Hospital, Taipei, Taiwan; Department of Mathematics, Courant Institute of Mathematical Sciences, New York University, New York, NY, US

**Keywords:** Variation of morphology, dynamic diffusion maps, nonstationary time series analysis, ROSELAND

## Abstract

**Objective:** Quantifying physiological dynamics from nonstationary time series for clinical decision-making is challenging, especially when comparing data across different subjects. We propose a solution, called Universal Coordinate (UC), and validate it using two real-world surgical databases, focusing on underutilized arterial blood pressure (ABP) signals.

**Method:** We apply a manifold learning algorithm, Dynamic Diffusion Maps (DDMap) under the novel UC, to quantify dynamics from nonstationary time series so that cross-subject comparison can be achieved. The method is demonstrated using ABP signal and validated with liver transplant and cardiovascular surgery databases, both containing clinical outcomes. Sensitivity analyses were conducted to assess robustness and identify optimal parameters.

**Results:** The UC solution is validated by significant correlations between the derived index and clinical outcomes. Sensitivity analyses confirm the algorithm’s stability and help optimize parameters.

**Conclusions:** UC enables across-subject comparison of ABP signals dynamics quantified by DDMap, and hence repurposes typically discarded ABP signals in the operating room and can be applied to other nonstationary biomedical signals in both hospital and homecare settings.

## 1 Introduction

Manifold learning is a versatile unsupervised machine learning technique used for visualization, dimension reduction, feature extraction and denoising. Recently, it has been applied to analyze nonstationary time series [1, 2], particularly multivariate ones, allowing phase space parametrization or approximation by a low dimensional manifold. Through manifold learning algorithms, complex dynamics of systems like brain activity [3] and respiration [4] can be extracted and quantified for tasks like visualization [5], prediction [2] and classification [3].

Dynamic diffusion map (DDMap) is a manifold learning algorithm specifically designed for nonstationary biomedical time series [1]. Based on *RObust and Scalable Embedding via LANdmark Diffusion* (ROSELAND) [6, 7], a computationally efficient variant of *Diffusion Map* (DM) [8], DDMap leverages the dataset’s spectral structure to quantify time series dynamics. This process is supported by the spectral embedding theorem [9, 10], recent convergence results [11, 12], and its robustness to noise [7, 13, 14]. The DDMap algorithm involves three main steps. First, follow a preassigned rule underpinning the dynamics to convert a continuous univariate nonstationary time series *y*_*t*_ ∈ *R, t* ∈ [0, *T*] into a discrete multivariate one *Y*_*k*_ ∈ *R*^*p*^, *k* = 1,…, *N* with a low-dimensional *phase space* modeled by a manifold, which we call *phase manifold*. Second, apply ROSELAND to suppress noise and embed *Y*_*k*_ into a low dimensional space, denoted as *Z*_*k*_ ∈ *R* ^*q*^, where *q* < *p* [7, 13, 14]. Third, quantify the system’s dynamics from *Z*_*k*_ with a suitable tool. DDMap has been successfully applied to study respiratory dynamics from electrocardiograms [4], predict short term liver transplant outcome [2, 15], and predict sleep stage from electroencephalogram (EEG) [3].

Despite its success, DDMap faces a critical challenge inherited from diffusion maps (DM) and ROSELAND: how to compare the embeddings of *different* point clouds, or databases, particularly those from different subjects? When applying DM or ROSELAND *separately* to multiple point clouds, it is unclear how to compare their embeddings due to differences in sampling distributions, supports, and imbalanced sample sizes (see Figure 1(b)). Existing methods like Procrustes alignment [16], α-normalization [8] and Nystrom extension [17] only partially address the issue, and time series analysis with DDMap is not an exception. The fundamental challenge of comparing embeddings from different datasets is crucial for analyzing time series dynamics across subjects.

**Figure 1.**
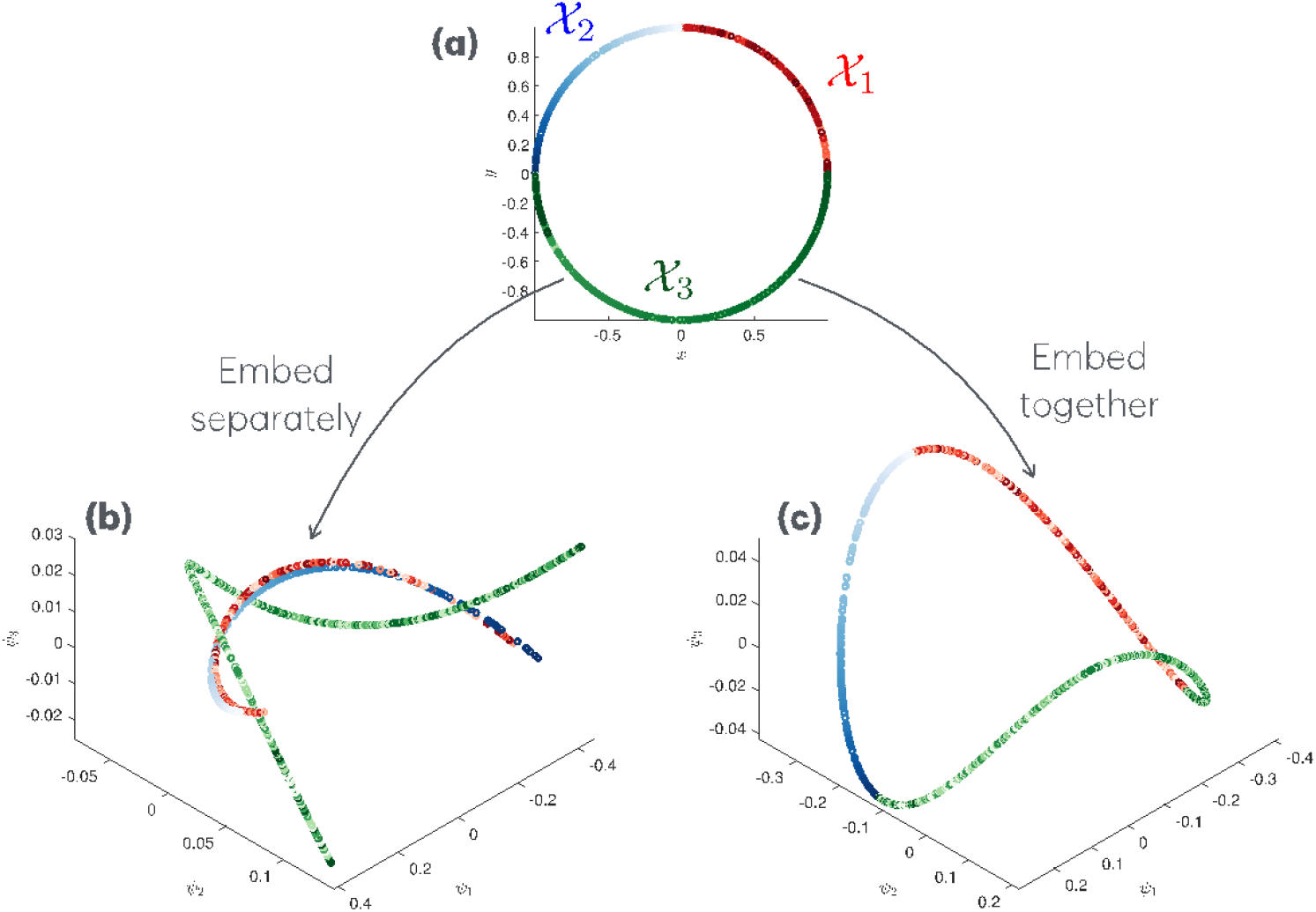
(a) Three point clouds simulate three time series from three subjects, marked by red, blue and green. The gradient color indicates the sampling time sequence. χ_1_ is a random but nonuniform sample. χ_2_ is a monotonically increasing sample. χ_3_ is an oscillation over π and 2π that repeats for π times. (b) The ROSELAND embeddings of χ_1_, χ_2_, and χ_3_ separately are marked in red, blue, and green separately, with the gradient color indicating the sampling time sequence. (c) The ROSELAND embedding of χ_1_ ∪ χ_2_ ∪ χ _3_, where we mark the embedded points with the associated color and gradient color.

To address this challenge, we propose a simple yet theoretically grounded framework called *universal coordinate (UC)*, built upon the concept of a *phase manifold*. UC is constructed by using a large reference dataset of the same type as the databases to be compared. This reference dataset is composed of multiple subjects with diverse physiological profiles but similar sensing status. Under certain assumptions that we will elaborate on below, the dataset serves as the ingredient for constructing the *phase manifold* that captures shared structure in physiological dynamics across subjects. After applying DDMap, this manifold provides a common coordinate system, called UC, for comparing nonstationary time series. Intuitively, the phase manifold hosts the dynamics of interest, the UC functions as a *coordinate* of the phase manifold, and DDMap is an algorithm that recovers UC from the dataset.

The rationale behind UC is twofold. First, while physiological dynamics vary across subjects, these variations are not arbitrary; rather, they are governed by fundamental principles that shape our bodily constitution, including genetic and environmental influences. These shared principles are captured and modeled by the phase manifold. Second, dynamical statuses that are rare in one subject may be common in others. This cross-subject variability is modeled by diffusion processes on the phase manifold, which account for differences in sampling distributions, supports, and sample sizes. UC represents the coordinate of the diffusion process on the phase manifold, providing a common basis for comparison across subjects. We argue that UC effectively “charts” the phase manifold, and hence the claimed comparison purpose. See Figure 1(c) for an illustration of UC, where the *S*^1^ structure is recovered, the geometric relationship between point clouds is preserved, enabling the design of comparable dynamic quantities.

We demonstrate the UC framework using two arterial blood pressure (ABP) datasets collected during liver transplant and open-heart cardiovascular surgeries (CVS). We further validate UC’s effectiveness through sensitivity analyses. The body’s response to complex surgical events is intricately reflected in cardiovascular dynamics, leading to subtle perturbations in evolving pulse cycles within the ABP waveform. Even minor stimuli can trigger complex responses, causing subtle variations in pulse cycles, which are challenging to detect and quantify by naked eyes. We have shown that DDMap can extract this delicate information [2, 28]. In this paper, we show that UC further enables a comparable quantification of these dynamics. Last but not the least, we conjecture that UC can be applied to other manifold learning algorithms, generalizing existing solutions like Procrustes alignment [16], α-normalization [8] or Nystrom extension [17], when comparing multiple databases is the mission.

The paper is organized as follows. Section 2 reviews ROSELAND and DDMap, gives a phenomenological model describing the intrinsic cardiovascular dynamic and its relationship with the ABP signal, and details the challenges of comparing different databases collected from different subjects. Section 3 presents our UC framework with theoretical support. Section 4 describes the databases used for numerical validation of UC. Results are shown in Section 5, and the paper concludes with a discussion in Section 6. Additional model details with theoretical support are provided in the online supplementary material.

## 2 ROSELAND, DDMap, and challenge

### 2.1 A review of ROSELAND

ROSELAND [6, 7] is a computationally efficient variant of DM [8]. DM is a widely used technique for characterizing the complex structure of high-dimensional point clouds and is supported by a rich theoretical foundation, particularly under the manifold setup (see [8-11, 13] for a nonexhaustive short list of available literature). Despite its broad applicability, the computational cost of DM can be substantial, primarily due to its reliance on eigenvalue decomposition (EVD).

A common strategy for reducing this computational burden is to sparsify the neighborhood graph by using either the *k* nearest neighbors (kNN) method, with a small *k* ∈ *N*, or an ε-ball neighborhood, with a small radius ε > 0. However, these approaches can be problematic for noisy datasets, in which identifying true neighbors based on observed distances alone is often challenging. To handle this issue, a solution via *landmark diffusion* called ROSELAN was proposed in [6, 7]. Below we quickly summarize the algorithm.

Suppose the given dataset is a point cloud χ = {*x*_1_, …, *x*_*n*_} in a metric space with the metric *d*. Select a landmark set *Y* = {*y*_1_, …, *y*_*m*_}⊂ *R*_*p*_, where *m* is smaller than *n*. The first step of ROSELAND is constructing an *n*×*m* affinity matrix *W* from χ and *Y*, such that

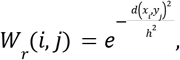

where *i* = 1, …, *n, j* = 1, …, *m*, and the metric *d* and bandwidth *h*> 0 are chosen by the user like those in DM. In practice it can be set as the 25% percentile of all pairwise distances between χ and *Y*, particularly when the data is noisy, according to current theoretical analysis [13]. Note that *W*_*r*_ (*i, j*) is the similarity between *x*_*i*_ and *y*_*j*_, so 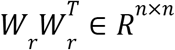 encodes similarities between points in χ. Next, we construct an *n*×*n* degree matrix *D*_*r*_, such that 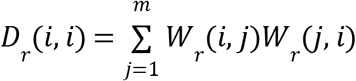, where *i* = 1, …, *n*. Denote the singular value decomposition (SVD) of 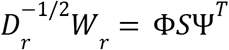 where *S* contains singular values 1 = *s*_1_ > *s*_2_ ≥ *s* _3_≥…*s*_*m*_ of 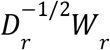 and Φ and Ψ are orthogonal matrices containing the corresponding left and right singular vectors, where the left singular vectors are ϕ_1_, ϕ_2_, …, ϕ_*n*_ ∈ *R*^*n*^. The SVD of 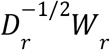 is directly related to EVD of 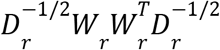, which is similar to 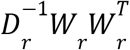. Clearly, 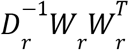 is a transition matrix on χ, which geometrically means that to diffuse from *x*_*i*_ to *x*_*j*_, we force the diffusion path to bypass the landmark set *Y*. Finally, the ROSELAND Map is defined as

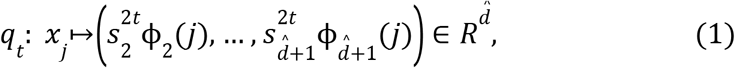

where *j* = 1, …, *n, t* > 0 is the user-defined *diffusion time*,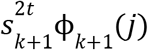 is the *k*-th *diffusion coordinate*, and 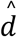 is the user-defined embedding dimension. In practice, while the parameter *t* can be determined by the von Neumann entropy [29], we simply set it to *t* = 1 following common practice. On the other hand, 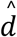 by the elbow method or profile likelihood [30]. The *ROSELAND Distance* between two points *x*_*i*_ and *x*_*j*_ is defined as

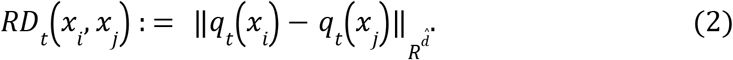

There are several benefits of ROSELAND. First, the computational complexity of RM is *O*(*nm*^2^), and in practice we take *m* = *n*^β^ and β≤1/2 so that *nm*^2^ < *n*^2^. Second, under the manifold setup, Roseland Map recovers the spectral embedding of the manifold. Third, ROSELAND is robust to heteroscedastic, colored and large noise, which is commonly encountered in practice. We postpone more theoretical details to Appendix A.1. For practitioners, the practical meaning of the spectral embedding and the robustness is that we obtain a low dimensional parametrization of the inaccessible manifold *M* even if the sample is noisy; that is, each high dimensional, possibly noisy, sample point can now be parametrized by a 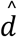 -dim vector that is less noisy. We could view the embedded points, 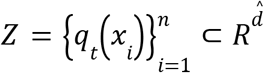, as a new dataset that is sampled from *practically* the same manifold that encodes the desired dynamical information and less impacted by noise.

### 2.2 A review of DDmap

DDMap [1] was designed based on ROSELAND to uncover subtle, previously unnoticed structures embedded in the signal. It shares properties of ROSELAND, for example, reconstruction of the global manifold structure with rigorous theoretical support. It is suitable for biomedical waveform signals like electrocardiogram (ECG) [4], photoplethysmography (PPG) [15], ABP [2, 28], EEG [3], etc. The algorithm is summarized below.

Given a recorded oscillatory biomedical waveform, denoted as *x* ∈ *R*^*N*^, where *N* indicates the number of sampling points, that is sampled at the sampling rate *f*_*s*_ Hz; that is, the recording lasts For *N*/*f* _*s*_ seconds. We could convert the univariate time series *x* into a point cloud 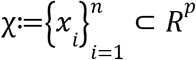, which is a multivariate time series, following a predesigned rule that is specific to the given time series. For oscillatory time series like PPG, ABP, or ECG, the rule can be “dividing the time series into cardiac cycles”; for EEG signals, the rule can be “the local power spectrum”. See Section 4.3 for the precise signal processing steps using ABP as an example. Then, apply ROSELAND on χ and obtain a 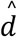-dim embedding of the waveform.

### 2.3 Main challenge and existing solutions

When DDMap is applied separately to datasets collected from different individuals, the resulting embeddings are generally not directly comparable. This limitation is inherited from ROSELAND (and DM) and stems from two main sources. First, RM is generally non-unique, partly due to the non-uniqueness of the SVD used in its construction. The singular vectors obtained from SVD are not uniquely determined and may vary under rotations within subspaces associated with repeated singular values. A similar ambiguity arises in DM, where the eigenvectors of the diffusion operator may also be non-unique, particularly when eigenvalues are repeated. Second, the RM depends on the sample size, sampling density, and support of the sampling distribution. These sources of variability are related to broader concepts in the literature, including distribution shift and, more specifically, support mismatch or heterogeneous support. Consequently, when multiple datasets have unequal sample sizes, substantially different sampling densities, or markedly different distribution supports, their resulting embeddings may not be directly comparable. Figure 1 illustrates this issue.

Several approaches have been proposed to mitigate this challenge. One simple strategy is to reduce complex time series to scalar summaries. For example, an ABP time series can be summarized by its systolic or diastolic blood pressures, yielding scalars that can be compared across subjects, albeit at the cost of losing information about the underlying dynamics. More sophisticated approaches include normalizing diffusion coordinates to unit variance and applying Procrustes alignment [16] to improve the comparability of embeddings. These methods can work well when the underlying point clouds have similar distribution supports and structures. When sampling densities differ, density-corrected kernels, such as the α-normalization scheme in DM [8], can help mitigate density-related effects, particularly when density heterogeneity is not too pronounced. Optimal transport is also an alternative technique that can be considered, if the domain of interest is known [31]. Another approach is the Nystrom extension [17], which uses an embedding constructed from one point cloud to extend the embedding to other datasets; this is most useful when the datasets have sufficiently compatible underlying structures and distributions. Once embeddings have been appropriately aligned, methods such as kernel fusion [18] or its generalizations can be used to quantify differences across datasets. However, these approaches generally rely on assumptions about structural compatibility or successful alignment that may not hold in general. Although each method addresses certain aspects of the problem, none fully resolves the combined challenges of embedding non-uniqueness, unequal sample sizes, heterogeneous sampling densities, and differing distribution supports.

More generally, when we analyze nonstationary time series using DDMap, we encounter the challenge of non-comparability, even if we assume that the dynamics of different subjects are the same. See Section 3.1 for detailed modeling information. In practice, the observed datasets may differ in length (sample size), in the frequency with which different dynamical states are observed (sampling density), and in the dynamics (distribution support). These differences, together with the non-uniqueness of the resulting embeddings, complicate direct comparisons across subjects. Because existing approaches do not adequately address all these challenges simultaneously, a new framework is needed to establish a common coordinate system for comparing nonstationary physiological dynamics across subjects.

## 3 Mathematical model and Universal coordinate

The mathematical model for studying nonstationary time series using DDMaps is given in Section 3.1. To handle the challenge mentioned in Section 2.3, we introduce our framework called *universal coordinate* (UC) as a solution in Section 3.2, where we give the guidelines for constructing UC in biomedical signal processing. The theoretical support is given in Section 3.3.

### 3.1 Manifold model for the cardiovascular state

We now introduce a mathematical model for the proposed framework. For simplicity, we focus on modeling cardiovascular status captured by ABP signals, although the general approach can be readily generalized to other suitable physiological signals. The ABP signal is recorded using a strain-gauge-based pressure transducer connected to a fluid-filled tubing system [32]. This transducer converts the mechanical pressure of the fluid into an electrical signal, which is digitalized the ABP signal that we analyze. The transducer mechanism suggests that each heart contraction and stroke can be conceptualized as an impulse applied to the cardiovascular system, with the resulting cardiac cycle in the ABP signal representing the system’s response. The resulting response, reflected in the ABP waveform, provides a surrogate measure of the underlying *cardiovascular system’s state (dynamics)*.

See Figure 2 for an illustration of this intuition. Mathematically, suppose the ABP signal recorded from the *i*th subject is denoted as *f* _*i*_ (*t*), the *k*-th cardiac stroke occurring at *t*_*k*_ (Figure 2(a)), and there are *m*_*i*_ cardiac cycles in the recording. In this case, the cardiac cycle in *f*_*i*_ after *t*_*k*_ can be viewed as a surrogate measure of the cardiovascular state. With a proper discretization, we obtain a high-dimensional time series

**Figure 2.**
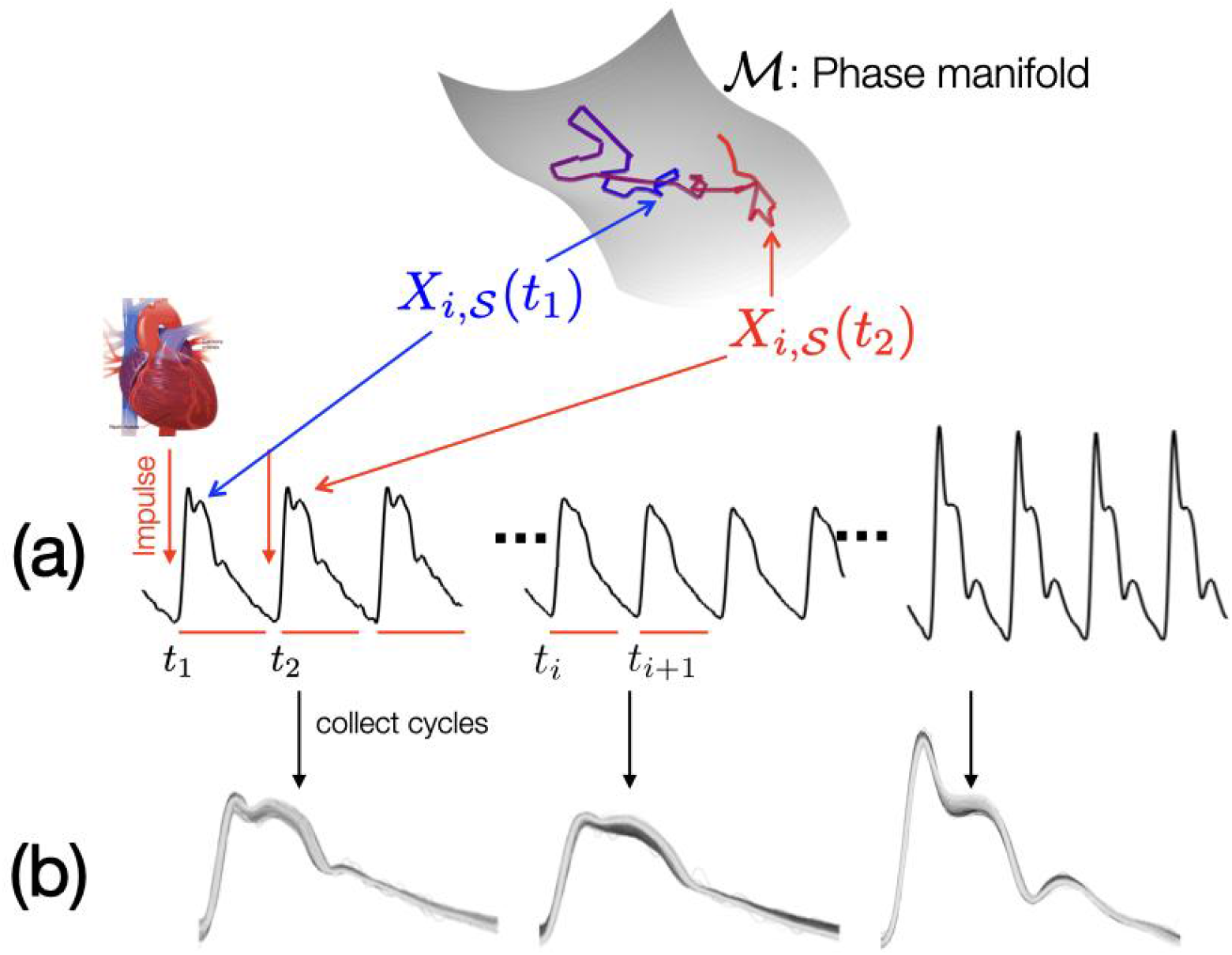
An illustration of the proposed model for the intrinsic cardiovascular dynamics and phase manifold, and its relationship with the ABP signal.

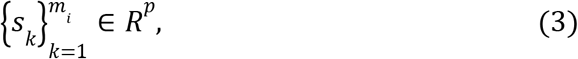

Where *s*_*k*_ is the *k*-th discretized cardiac cycle (Figure 2(b)). This conversion from the univariate ABP signal to discrete multivariate cardiovascular state time series allows us to further explore the cardiovascular state dynamics.

Despite the simplicity of this intuition, however, physiological processes arise from complex interactions between genetic predispositions and environmental influences, making it difficult to fully characterize the cardiovascular status using conventional differential equations. Rather than attempting to formulate a comprehensive mechanistic model, we represent cardiovascular dynamics using a stochastic differential equation (SDE) evolving on a low-dimensional, compact, and smooth manifold *M* isometrically embedded in *R*^*p*^ via ι which we refer to as the *phase manifold*. We do not assume that a specific parametrization of *M* is known a priori.

For the *i*-th subject, where *i* = 1, 2, …*m*, consider the following SDE to model its cardiovascular status:

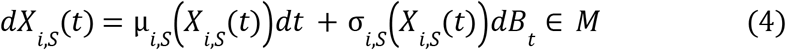

with the initial status *X*_*i,S*_ (0)∈*M*, where *S* is the *sensing status, X*_*i,S*_ (*t*) is the *cardiovascular state* at time *t*, µ_*i,S*_ : *M*→*TM* is the drifting term, σ_*i,S*_ : *M*→*TM*⨂*TM*_*_ is the diffusion term, and *B*_*t*_ is the standard Brownian motion on *M*. Here, the sensing status *S* includes the subject’s health status, the sensor’s quality, the environment, etc. We could further model the sensing status when necessary. Since for our application we could reasonably assume the sensing status is the same across subjects, for simplicity, we will not further quantify this dependence and unless otherwise noted, we suppress the dependence on *S* in notations to streamline the presentation. We emphasize that the proposed model is phenomenological; that is, rather than providing a detailed mechanistic account of the underlying physiological processes, it offers a simplified yet sufficiently informative representation of the system’s overall dynamics.

Assume µ_*i*_ and σ_*i*_ are non-degenerate and satisfy the Lipchitz condition so that a unique strong solution of SDE exists, and the solution is continuous. It is important to underscore that while the drift and diffusion terms depend on subjects, the dynamics *all* live in *M*. In other words, we model the cardiovascular state as a continuous diffusion process on the phase manifold.

Then, we sample *X*, potentially nonuniformly, and form a multivariate time series as

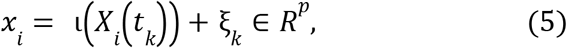

where *t*_*k*_ represents the timing of the *k*-th cardiac stroke and ξ_*k*_ accounts for inevitable noise. For simplicity, we assume ξ_*k*_ is stationary with finite fourth moments. When the ABP signal is noise free, 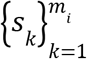 in (3) constructed from the ABP signal is a the clean sampled cardiovascular state 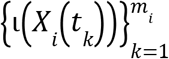, while in practice it the noisy data 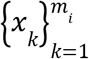 in (5).

### 3.2 Universal coordinate (UC)

Constructing UC is straightforward under the dynamic model given in Section 3.1. See Figure 3 for an illustration of the overall UC solution. We collect ABP signals from *n* different subjects with *similar* sensing status as possible. For the ABP signal 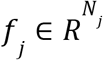 recorded from the *j*-th subject, build a point cloud embedded in the *p*-dim Euclidean space with *m*_*j*_ cycles, denoted as χ_*j*_ (the detailed numerical implementation is postponed to Section 4.3). We refer to a set

**Figure 3.**
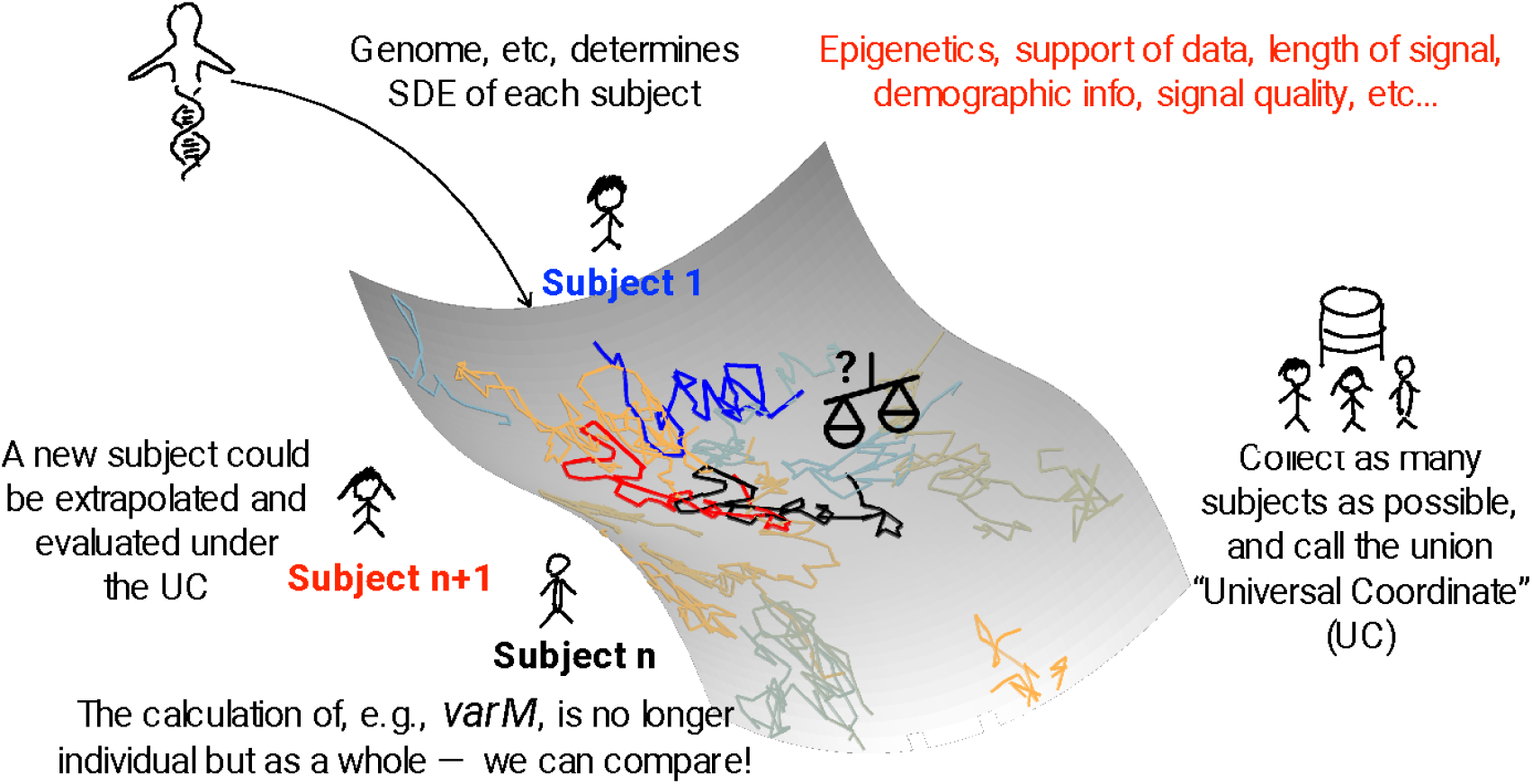
The overall flowchart of the proposed UC framework. By collecting *n* subjects, we construct the UC that charts the phase manifold that hosts the dynamics of different subjects with similar sensing status. The (*n* + 1) -th subject indicated by red color, could be merged to the UC for the dynamic’s evaluation. With the UC, the dynamics of different subjects is therefore comparable.

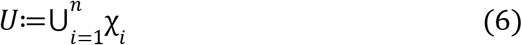

as the *pooled dataset*.

Take the (*n* + 1)-th subject. Denote its associated point cloud as χ_*n*+1_ that contains, and run DDMap on *U* ∪ χ _*n*+1_ so that χ _*n*+1_ is embedded in 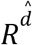 together with *U*. Suppose there are 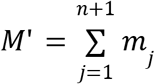 points in *U* ∪ χ. Denote the DDMap of *U* ∪ χ_*n*+1_ as

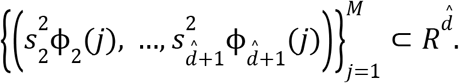

The UC for the cardiovascular state dynamics of the *k*-th sample of the *i*-th subject is 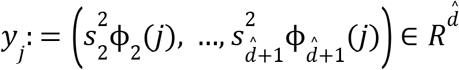, where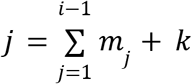. Clearly, 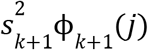 is the *k*-th component of the UC.

The UC allows us to explore the dynamics of the subject of interest by exploring the dynamics of χ _*n*+1_ via this embedding. The term “coordinate” in UC literally means constructing the coordinate of the phase manifold, which is achieved by gathering ample data that represents various inherent inter-individual variabilities, enabling meaningful comparisons between different subjects.

At a high level, UC *standardizes* DDMap in the following sense. As is discussed in Section 2.3, due to uneven sample sizes, skewed distributions, and uneven supports, DDMaps of different χ_*i*_ are generally not comparable. UC mitigates this comparability issue via collecting different subjects that encodes potential variabilities between subjects. Indeed, with sufficient collection of subjects, the UC well charts the phase manifold. As discussed in Section 3.3 below, with some assumptions, an analysis with UC is less impacted by the individual variation and hence provides a standardization. It is also worth noting that the UC concept can be applied to other datasets beyond time series, which will be reported in a future study.

### 3.3 Assumptions and consequent properties of UC

In practice, for each subject whose cardiovascular dynamics are governed by the SDE in (2), only one or a few sample trajectories are available, each starting from its own initial condition. To characterize the long-term behavior of these trajectories and facilitate comparisons across subjects, we assume that the phase manifold *M* is smooth, compact, and without boundary. Under our assumption, the time-homogeneous SDE of the *i*-th subject becomes positive Harris recurrent [33], with the invariance measure π_*i*_. In general, π_*i*_ depends on the subject-specific drift µ_*i*_ and diffusion σ_*i*_. Under this setup, sufficiently long trajectories exhibit ergodic behavior, such that time averages of suitable observables converge almost surely to their expectations with respect to π_*i*_ [33]. Mathematically, if we denote 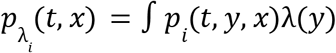 to be the marginal density of the *i*-th subject’s SDE at time *t* > 0, where *p* _*i*_ (*t, y, x*) is the transition density and λ_*i*_ is the initial measure of the *i*-th subject, then we have exponential ergodic convergence 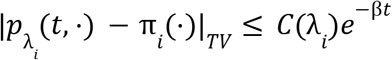 for some β > 0, where TV means total variation. In this sense, the long-term statistical behavior of each trajectory is characterized by its associated invariant measure. These invariant measures provide a principled basis for quantitatively characterizing and comparing cardiovascular dynamics across subjects.

The main challenge is that the support of π_*i*_ might be a strict subset of *M*, nonuniform over its support, and vary from subject to subject due to the inter-individual variability. In other words, certain regions of *M* may be infrequently visited by one subject’s dynamic trajectory, yet regularly traversed by another, so that each subject would provide *partial observation* of the phase manifold. Central to our methodology is integrating *partial observation* to obtain a full exploration of the phase manifold.

Since we assume the driving Brownian motions and initial distributions of different subjects are mutually independent, the associated diffusion processes are mutually independent as well. The resulting invariant measures {π_*i*_} _*i*=1,…,*n*_ of *n* different subjects can be viewed as i.i.d. sampled from a measure-valued random variable *Z*: (Ω, *F, P*) → *P*(*M*), where (Ω, *F, P*) is the probability space and *P*(*M*) is the set of probability density functions on *M*. Denote *T*_*i*_ : = γ_*i*_ *T* to be the observation time of the *i*-th subject, where *T* > 0, γ_*i*_ ∈ (0, 1) so that 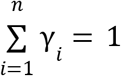 is the ratio of observation times of different subjects. Then, asymptotically when *T* → ∞, the pooled dataset (or total mixture) on *M* converges almost surely to the time-weighted mixture of their individual invariant measures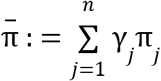. We assume that the density function of 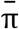, denoted as 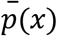, is sufficiently uniform; that is, 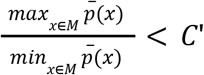 for a constant *C*’≥1. This assumption says that by pooling together all recordings, we obtain a well-behaved point cloud over the phase manifold so that the consequence algorithm, DDMap, can work well.

## 4 Material and statistics

We performed a clinical data validation and the sensitivity analysis to demonstrate the effect of the proposed UC framework using two ABP datasets from diverse medical conditions, including liver transplant surgery and CVS. Both datasets involve critically ill patients.

### 4.1 Liver transplant database

Liver transplant surgery is the only option for some patients with severe liver disease, with success largely measured by recipient survival within three months post-surgery and laboratory indicators of liver graft function. We found that the morphological variability of ABP waveforms, quantified by *variability of morphology* (*varM*), during the neohepatic phase, preceding wound closure and marking initial liver graft function, encodes clinically useful information; that is, *varM* correlates with presurgical liver condition and post-surgery liver function [2]. Previous studies [2, 5] show that *varM* is more closely associated with clinical outcomes than traditional indices derived from ABP, such as systolic blood pressure, diastolic blood pressure, or blood pressure variability.

Data were collected from a single-center prospective observational study adhering to the 1975 Declaration of Helsinki, with Institutional Review Board approval (IRB No.: 2017-12-003CC and 2020-08-005A) and written informed consent. ABP waveform data, essential for this surgery, were recorded throughout the procedure using the patient monitor (GE CARESCAPETM B850, GE Healthcare, Chicago, IL) with S5 Collect (GE Healthcare) software at a 300 Hz sampling rate. Clinical scores included MELD-Na, assessing liver disease severity before surgery, and L-GrAFT10 score, predicting post-surgery graft failure. The database contains 85 cases, encompassing diverse liver disease conditions such as acute liver failure or liver cancer. In this study, we use presurgical ABP waveforms to construct UC.

### 4.2 Cardiac surgery database

Inspired by the liver transplant surgery analysis, we hypothesize that for the CSV dataset, *varM* evaluated before anesthesia correlates with baseline preoperative renal function. The CVS database was collected from a single-center prospective observational study conformed to the 1975 Declaration of Helsinki, with Institutional Review Board approval (Shin Kong Wu Ho Su Memorial Hospital IRB No 20181205R) and informed consent. Physiological waveform data were collected using the IntelliVue MX800 patient monitor (Philips, Amsterdam, Netherlands) and ixTrend software (ixellence GmbH, Wildau, Germany) at a 125 Hz sampling rate. The database contains 107 cases. The UC is constructed from pre-anesthesia data, reflecting baseline conditions, with kidney function quantified by the estimated glomerular filtration rate (eGFR) from preoperative laboratory measurements and demographic variables.

### 4.3 Converting ABP signal into discrete-time multivariate time series

In this subsection we detail the numerical implementation of the multivariate time series described in (3). The sampling rate of ABP is *f*_*s*_ = 300. The fiducial point as the starting point of a cardiac cycle is the maximum of the first derivation of its ascending part. Segment the ABP signal *x*_*i*_ ∈ *R* ^*n*^ recorded from the *i*-th subject into cardiac cycles and denote *n* _*i,k*_ as the *k*-th landmark for the starting point of the *k*-th cycle and assume there are *m*_*i*_ cycles in *x*_*i*_. The *k*-th pulse cycle is *x*_*i,k*_ *∶* = [*x*_*i*_(*n* _*i,k*_,) *x*_*i*_ (*n*_*i,k*_ + 1), …, *x*_*i*_ (*n*_*i,k*_ + *q* – 1)]^*T*^, where *p* is the minimal length of all segments to which each pulse is truncated to ensure a uniform size. The point cloud is thus 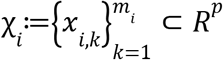, which is the desired multivariate time series. The UC *U* is then constructed from a pool of subjects with the same sensing status using DDMap.

### 4.4 Variation of Morphology (*varM*) index

Take the embedding dimension 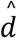; that is, DDMap converts a pulse cycle into a vector of length 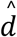. The elbow method suggests 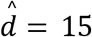, since the eigenvalues decay rapidly so that 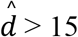 eigenvalues can simply be neglected. With the UC of dimension 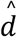, we quantify the dynamics in the following way [2], which can be viewed as a layman’s implementation of the kernel estimate of the drifting term [19]. In order to reduce the impact of noise, for the *i*-th subject, a moving median filter followed by a moving mean filter is first applied:

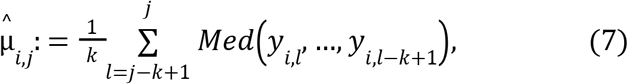

where *y*_*i,l*_ is the UC of the *l*-th pulse cycle of the *i*-th subject. Both filters are with a window length *k*. Then, for the *i*-th subject, calculate

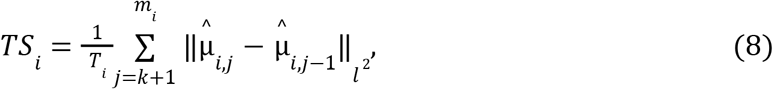

where *T*_*i*_ is the length of recording. For convenience in future clinical applications, we convert *TS*_*i*_ to the *variation of morphology index* (*varM*), which is within the 0-100 range, by

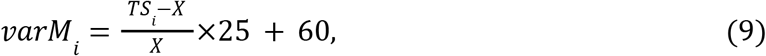

where *X* is determined from the whole dataset. Note that the considered *varM* captures the “traveling distance” of the dynamics driven by the drifting term, since 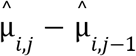 is essentially a noisy estimate of the drift term of the *i*-th subject at time *n*_*i,j*−1_.

### 4.4. Validation of the proposed UC

Our validation of whether the UC improves performance of *varM* is entirely data-driven and supported by clinical information. For a database of *n* recordings and 0 < *m* < *n*, we explore how the pooled dataset size impacts stable UC construction by randomly sampling *m* recordings to form UC for *Q* ∈ *N* times and reporting the median and interquartile range (IQR) of the Spearman’s correlation coefficients (SCC) between *varM* and the clinical indices of interest over these *Q* times random samples. It is critical to note that when *m* = 0, it means we evaluate *varM* without using UC.

We also evaluate which type of dataset is more effective for UC construction by comparing UCs from different surgical phases or datasets. Additionally, we test the sensitivity of key parameters, including the embedded dimension of DDMap (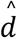 in (1)) and the smoothing window length (*k* in (7)), where for 1≤*m* < *n* yielding median performance, we resample *m* recordings *Q* times and report the median and IQR of SCC between the *varM* and clinical indices.

During the sensitivity analysis, we vary one parameter at a time while keeping the others fixed at their default values. The default parameters for *varM* calculation are 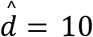, *m* = 40, and *K* = 21, based on our previous work [2]. Finally, we examine whether DDMap is crucial for quantifying dynamics with UC. We run an alternative experiment with exactly the same procedure except DDMap is not applied; that is, we calculate *varM* using (7)-(9) with the pooled dataset (6) instead of UC.

Waveform data processing and the DDMap algorithm, based on ROSELAND, were implemented in C# (v.12.0, Microsoft) with the LAPACK library (v.3.8.0, Linear Algebra PACKage team). Statistical analyses were conducted using R (version 4.4.0, R Core Team).

## 5 Results

We first illustrate the impact of UC in Figure 4. Also recall Figure 1 with the simulated data. The UC is constructed from 107 cases in the baseline period of the cardiac surgery (in total 38,180 pulses represented as grey dots in the lower panel). The demonstrated two cases have different clinical backgrounds and we expect to see a difference between their ABP morphology. They have similar blood pressure ranges, but it is not clear how different their morphologies are. It is easy to see the difference between the DDMap embeddings of two cases with UC and those without UC; that is, we embed each case separately. This difference is particularly visible when we inspect the fourth, fifth and sixth eigenvectors. Specifically, without UC, while we could see two clusters of point clouds in the first 3 diffusion coordinates, the 4^th^, 5^th^ and 6^th^ diffusion coordinates lack separation and exhibit excessive twisting.

**Figure 4.**
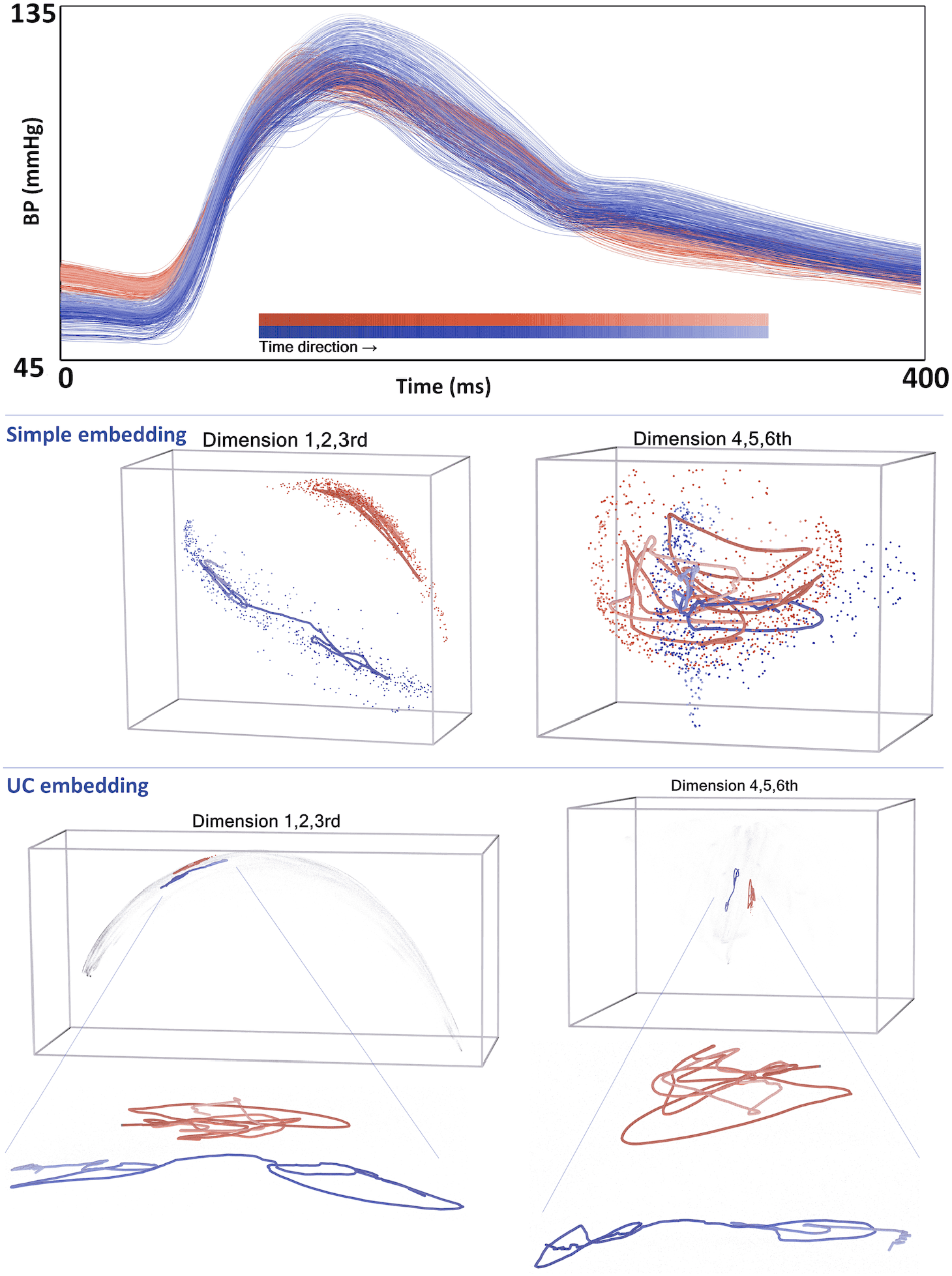
Demonstration of UC using two ABP waveforms of two cases from the neohepatic phase of liver transplant data (Case Blue: 575 pulses, Case Red: 785 pulses). Faded color indicates time sequence. Despite similar blood pressure ranges (upper panel), *varM* are different. Without UC, the first 3 diffusion coordinates show some clustering, where thick lines indicate their trends (middle panel), but the 4^th^, 5^th^, and 6^th^ diffusion coordinates lack separation and exhibit excessive twisting. With UC, both cases are better separated across the first 6 diffusion coordinates.

Below are the sensitivity analyses results. *U*_*c,m*_ (resp. *U*_*l,m*_) denote UCs constructed from *m* recordings from the CVS database before anesthesia as the baseline (resp. *m* recordings from the liver transplant surgery database during the pre-surgical phase as the baseline).

### 5.1. Liver transplant dataset

We report the SCC between *varM* during the second half of the neohepatic phase of liver transplant surgery and various clinical indices. The MELD-Na score (SCC=-0.437, p=0.00004) reflects pre-surgery liver disease severity, and the L-GrAFT10 score (SCC=-0.288, p=0.0085) assesses the risk of post-surgery graft failure. Since both indices are low and *varM* is high in better clinical conditions, a negative correlation with *varM* is expected. We used *m* = 0, 5, 10, 15, 20, 30, 40, *K* = 11, 15, 21, 25, 31, and *Q* = 60. See Figure 5 for the result.

**Figure 5.**
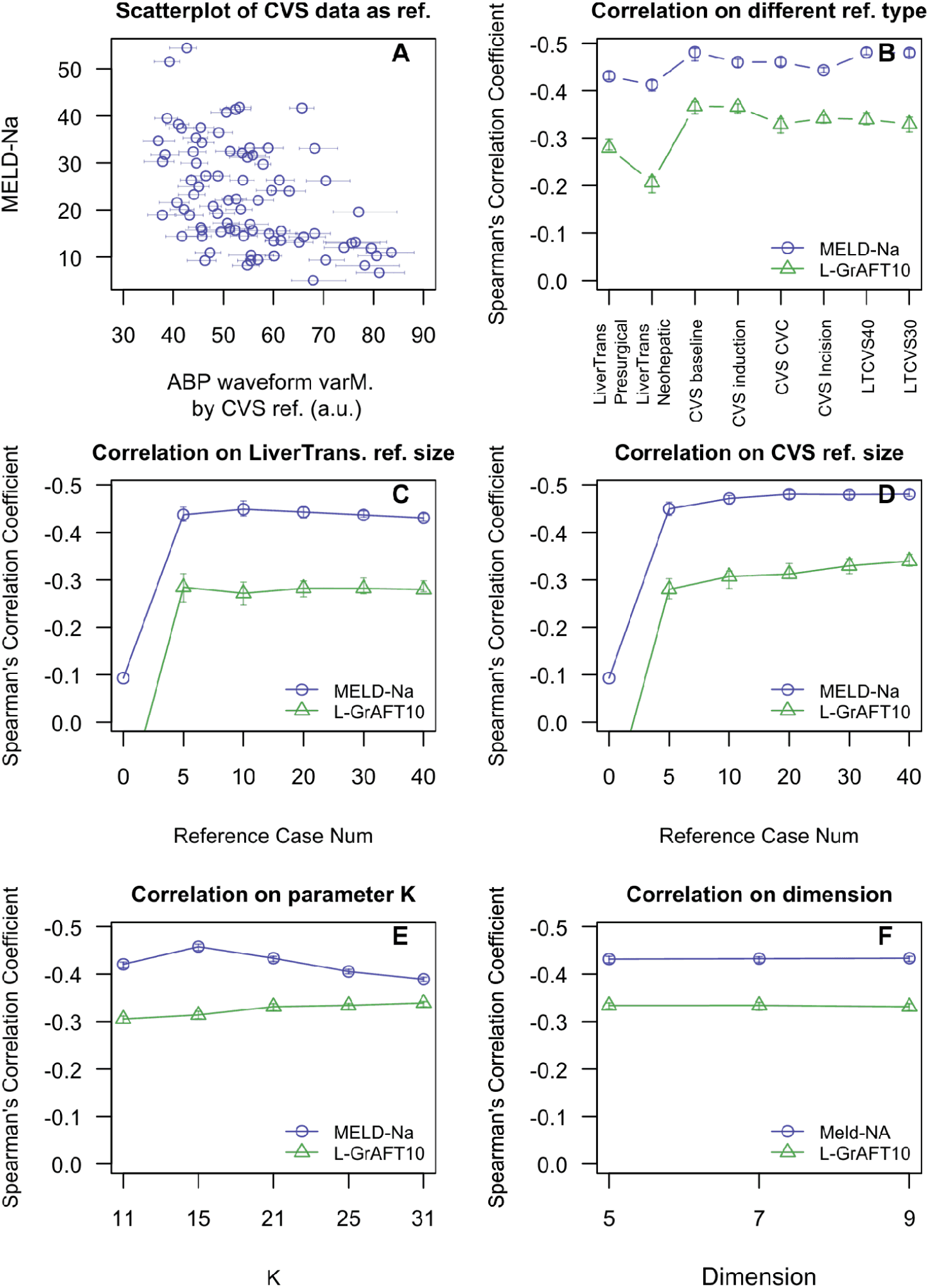
Sensitivity analysis on the liver transplant surgery database, focusing on the association between *varM* and clinical conditions, including Meld-Na score representing pre-surgery liver disease severity, and L-GrAFT10 score representing the risk of post-surgery live graft failure. A more negative correlation with *varM* indicates a better association. Error bars represent the interquartile ranges.

The results show three key findings. First, utilizing UC yields a significantly more negative SCC compared to when UC is not used. Second, UC performance varies based on pooled dataset size, source, and clinical outcome. CVS data from several surgical steps (pre-anesthesia, anesthetic induction, central vein cannulation, or surgical incision) as the pooled dataset yields overall better results than that from the liver transplant dataset. Mixing liver transplant and CVS data offers slightly better results. When using *U*_*c,m*_, increasing *m* enhances the association between the MELD-Na score and *varM*, a trend not seen with *U*_*l,m*_. Both MELD-Na and L-GrAFT10 scores slightly favor *U*_*c,m*_. Third, the embedding dimension 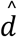 is relatively stable, with MELD-Na slightly favoring higher 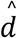, though this is not observed with L-GrAFT10. The smoothing window length *K* is also robust, with different *K* values providing similar associations, though the optimal *K* differs for MELD-Na and L-GrAFT10.

### 5.2 Cardiovascular surgery dataset

We evaluate the correlation between *varM* calculated with *U*_*c,m*_ and the baseline renal function, represented by eGFR, before anesthesia. Since higher eGFR and higher *varM* indicate better conditions, a more positive correlation suggests a better result. We set *m* = 0, 5, 10, 15, 20, 30, 40, 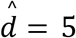, and *Q* = 60. The results are summarized in Figure 6.

**Figure 6.**
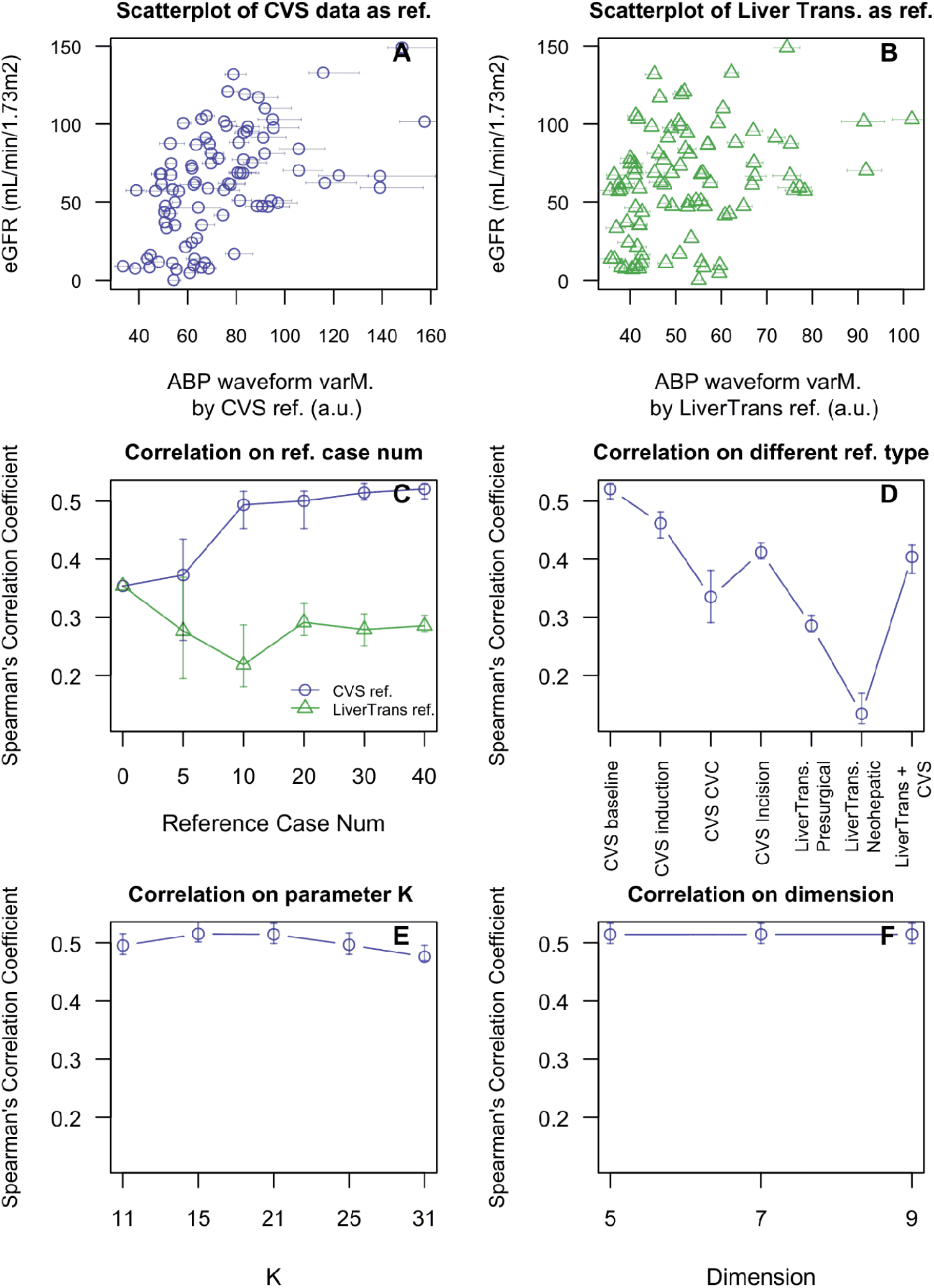
Validation with the CVS dataset, showing the correlation between presurgical renal function (eGFR) and *varM* before anesthetic induction. A positive correlation is expected. Error bars represent interquartile ranges.

We have three main findings. First, with *U*_*c,m*_, SCC gradually increases, and the IQR consistently shrinks as the number of recordings increases from 0 to 60. This trend is less obvious with *U*_*l,m*_ (middle top panels in Figure 6). Second, using *U*_*c,m*_, we obtain the best performance compared to using CVS data from other surgical phases, including anesthetic induction, central venous cannulation. All of these results outperform using *U*_*l,m*_ or the data from the neohepatic phase as the reference. A mix of liver transplant and CSV datasets falls in between. Third, the choice of embedding dimension and smoothing window length is relatively robust. When 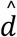, *m* = 40, and *K* = 21, there is a statistically significant correlation between *varM* and eGFR (SCC=0.525, p<10^−7^). Additionally, the scatterplots in Figure 4 visually demonstrate a stronger association between *varM* and eGFR with *U*_*c,m*_.

### 5.3 With or without DDMap?

We show evidence of the necessity of DDMap for the computation of *varM*. See Fig. 7 for the result. In both liver transplant and CVS datasets, the best correlation between the clinical index of interest and *varM* comes from using *U*_*c,m*_. The mix of *U*_*c,m*_ and *U*_*l,m*_ gives a close 2^nd^ best result, and the result is worst when DDMap is not included. The margin of the performance gain is largest in the CVS dataset when *U*_*c,m*_ is utilized. In the liver transplant dataset, the margin of the performance gain is larger when L-GrAFT10 considered compared with that of MELD-Na. In general, *varM* computation without DDMap presents a less clinical association but a narrower interquartile range. It deserves to mention that without DDMap, we face computational burden with more than 10 times computation time compared with that with DDMap.

**Figure 7.**
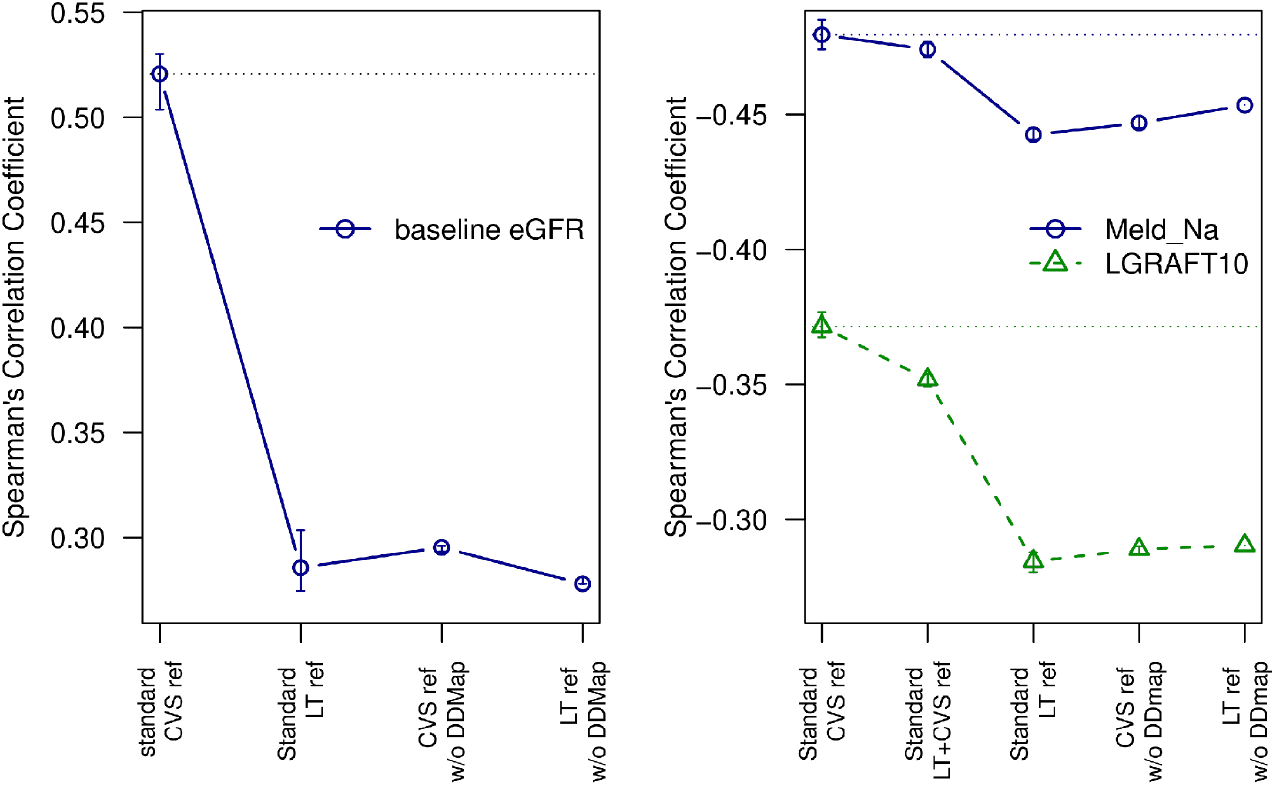
A comparison of *varM* constructed with and without DDMap under different UCs, where the correlation between *varM* and baseline renal function (resp. conditions) in the CVS (resp. liver transplant) database is shown on the left (resp. right) panel.

## 6 Discussion

Studying intricate physiological dynamics from nonstationary time series is challenging. While DDMap, an unsupervised manifold learning algorithm, has been successfully applied to various biomedical signals [2-4, 15], it often faces the challenge of how to compare different subjects. This paper introduces the UC framework to address this issue. By focusing on ABP waveforms, we show UC’s applicability using two real-world surgical databases and conduct sensitivity analyses. Our results validate UC’s usability and show that *varM* is robust to parameter choice. Additionally, we present a phenomenological model that captures essential cardiovascular state dynamics without digging into unknown mechanisms, and providing insights into general trends and behaviors, and connect it in a novel way to analyze ABP signal.

Enlarging pooled dataset enhances the association of *varM* with clinical indices, with significant improvement initially, which plateaus after 20 recordings (cases). This effect is most evident when using *U*_*c,m*_ for evaluation in the CVS database. Our findings suggest that while a large database is beneficial, the proposed algorithm remains effective even with limited data. On the other hand, the quality of UC construction depends on the chosen pooled datasets. CVS patients present diverse cardiovascular conditions, such as valvular heart disease, atherosclerosis, and coronary artery disease, leading to varied CVS waveform data. This diversity enhances UC’s ability to achieve a comparable *varM*. In contrast, liver transplant patients often exhibit a unique cardiovascular profile [20-22] known as hyperdynamic status, characterized by high cardiac output, hypervolemia, and low systemic vascular resistance. This results in a less diverse but more focused pooled dataset for UC construction, reflecting our validation results that suggest liver transplant pooled dataset have a “context-sensitive” effect. Combining pooled datasets from both surgical databases improves clinical associations, demonstrating robustness from both quantity and quality effects. Our analysis suggests the advantage of using the CVS dataset for UC construction in both surgical datasets.

Sensitivity analyses on the embedding dimension 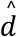 show limited performance gains at higher dimensions, indicating robustness and favoring computational efficiency. Computation of *varM* on hundreds of dimensions with DDMap is not only time-consuming but also ineffective. The *K* sensitivity analyses reveal some robustness across the two clinical databases, though different clinical outcomes exhibit varying *K* shapes, suggesting a complex relationship between *varM*, time scale, and clinical outcomes. While the clinical behavior and mechanisms of *varM* are becoming clearer, further studies are needed. Lastly, comparing the algorithm with and without the DDMap highlights DDMap’s crucial role in processing large datasets. These analyses suggest that the proposed UC framework is relatively robust to parameter choice, and it is recommended to apply DDMap to clean up the dataset and reduce its dimension before evaluating *varM*.

In biomedicine, comparison challenges arise due to inevitable inter-individual variability. For instance, comparing ABP recordings across subjects is difficult due to influences like vascular tone, cardiac contractility, and preload. Common approaches include converting ABP signals into scalar quantities, such as systolic or diastolic blood pressure, heart rate, or their variabilities [23], or extracting numerous features, like million [24], to gather all possible information. However, these methods risk either losing information through dimension reduction or overfitting with excessive features. The proposed UC framework offers a balanced solution, preserving as much information as possible while maintaining a manageable dimensionality.

The correlation between ABP waveform morphology and clinical status is supported by decades of medical knowledge. The association between *varM* and pre-operative status in both databases reflects the underlying clinical condition. In clinical practice, pooled data guide management and risk stratification based on the distribution of the general population. For example, recommended mean blood pressure for perioperative care is derived from prior data analysis. *varM* provides insights into individual patients’ conditions during operations, and standardized quantitative *varM* could enhance precision of decision making via condition assessment, risk stratification, and outcome prediction [27].

This study has limitations. First, *varM* is a newly established index, and there does not exist established ideal correlation values in our validation analyses to benchmark our results. While there are several publicly available databases, usually they do not have sufficient clinical information for our purpose. So prospective collection of new databases is necessary. Additionally, we only explored datasets from liver transplant surgery and CVS; future research should investigate other clinical databases. Moreover, our recordings span approximately 10 hours. While theoretically it is feasible, it remains to be validated if this approach is suitable for longer recordings over days or years, and what happens if we have much more subjects with much shorter recordings.

## 7 conclusion

The benefit of applying the proposed UC framework for quantifying physiological dynamics is supported by the significant correlation between the derived index *varM* and clinical outcome in two databases under consideration. We conclude that this technique has the potential to repurpose often ignored detailed morphology in ABP signals in the operation room for clinical decision making. Given the nature and flexibility of the UC framework, we anticipate that the algorithm could be applied to other signals in both hospital and homecare environments.

## Data Availability

All data produced in the present study are available upon reasonable request to the authors

## ACKNOWLEDGMENTS

This work was supported in part by the National Science and Technology Council, Taiwan (NSTC 112-2115-M-075-001 and 112-2314-B-075-070-MY2).

## A. APPENDICES

### A.1 Properties of ROSELAND: spectral embedding and robustness to noise

Suppose we have sampled *n* points 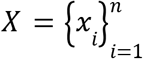 identically and independently (i.i.d.) from *M* following a density function *p*_*x*_(*x*) that is smooth with *p*_*x*_(*x*) > 0. While we have these samples, we do not know how the manifold is parametrized or even its dimension. The only information we have is that these samples are located on a low dimensional and smooth manifold; that is, we cannot access the manifold but only the high dimensional sample points.

Also, we sample *m* points i.i.d. from *M*, denoted as 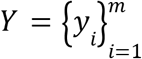, which is sampled independent of *X*, following another density function *p*_*y*_ (*x*) that is smooth with *p*_*y*_ (*x*) > 0.

Under these assumptions, it is shown in [7] that when some mild assumptions of the bandwidth are fulfilled, when *n* is sufficiently large and the bandwidth *h* in (3) is chosen properly, the RM approximates with high probability the *spectral embedding* of the manifold *M* [25]. Moreover, for any given small ε > 0, when the diffusion time is sufficiently small and 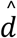 in (4) is sufficiently large, we obtain a spectral embedding that is deviated from being almost isometric with the deviation controlled by ε [9]. Mathematically, the top 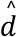 singular values and singular vectors in (2) coming from ROSELAND would approximate with high probability the top 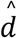 eigenvalues and eigenfunctions of the Laplace-Beltrami operator, and it has been well known that the manifold can be almost isometrically parametrized by sufficiently many top eigenvalues and eigenfunctions of the Laplace-Beltrami operator.

On the other hand, it is shown in [7] that ROSELAND is robust to independent large and high dimensional noise, which can be colored and heterogeneous. Colorness means the noise may have a dependent structure in each sample; heterogeneity means the noisy has different properties from sample to sample, including the covariance structure, tail behavior, etc. By largeness, we mean that ROSELAND can tolerate noise with the energy (the trace of the covariance matrix) that blows up at the rate slightly slower than 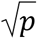, where *p* = *p*(*n*) is the dimension of the data so that 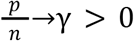 when *n*→∞. By combining these facts, RM could robustly and almost isometrically recover the phase manifold.

